# Temporal Drift in the Semantic Meaning of Pediatric Anxiety Terms in Electronic Healthcare Records

**DOI:** 10.1101/2025.03.09.25323626

**Authors:** Jordan Tschida, Mayanka Chandrashekar, Heidi A. Hanson, Ian Goethert, Surbhi Bhatnagar, Daniel Santel, John Pestian, Jeffery R. Strawn, Tracy Glauser, Anuj J. Kapadia, Greeshma A. Agasthya

## Abstract

**Objective:** To identify and measure semantic drift (i.e., the change in semantic meaning over time) in expert-provided anxiety-related (AR) terminology and compare it to other common electronic health record (EHR) vocabulary in longitudinal clinical notes.

**Methods:** Computational methods were used to investigate semantic drift in a pediatric clinical note corpus from 2009 to 2022. First, we measured the semantic drift of a word using the similarity of temporal word embeddings. Second, we analyzed how a word’s contextual meaning evolved over successive years by examining its nearest neighbors. Third, we investigated the Laws of Semantic Change to measure frequency and polysemy. Words were categorized as AR or common EHR vocabulary.

**Results:** 98% of the AR terminology maintained a cosine similarity score of 0.00 – 0.50; at least 90% of common EHR vocabulary maintained a cosine similarity score of 0.00 – 0.25. Laws of Semantic Change indicated that frequently occurring vocabulary words remained contextually stable (Frequency Coefficient = 0.04); however, words with multiple meanings, such as abbreviations, did not show the same stability (Polysemy Coefficient = 0.630). The semantic change over time within the AR terminology was slower on average than the semantic change within the common EHR vocabulary (Type Coefficient = -0.179); this was further validated by interacting the year and Type (Coef = -0.09 – -0.523).

**Conclusions:** The semantic meaning of anxiety terms remains stable within our dataset, indicating slower overall semantic drift compared to common EHR vocabulary. However, failure to capture nuanced changes may impact the accuracy and reliability of clinical decision support systems over time.

## 1 Introduction

There has been growing interest in using machine learning (ML) algorithms for clinical decision support systems (CDSS) [1]. ML for CDSS can revolutionize healthcare; however, the success of these applications depends on the quality of the data provided to the models. As more institutions begin to leverage natural language processing (NLP) techniques for CDSS, tools for determining when semantic drift emerges and quantifying the magnitude of the change can inform future algorithm development and enhance the accuracy of automated CDSS [2–5].

CDSS are especially important for diagnosing complex or challenging conditions, such as childhood anxiety. Anxiety disorders affect 32% of the pediatric population [6, 7], making the development of effective diagnosis and treatment strategies a high priority for mental health patients and providers [8]. CDSS could assist in monitoring a patient’s anxiety symptoms over time and alert clinicians of drastic or sudden changes in a patient’s condition. Early and accurate diagnosing of pediatric anxiety mental health disorder is important, as children with undiagnosed anxiety may develop reduced or delayed social skills, which can adversely affect their adult life [9].

In a clinical setting, the language that clinicians use to describe mental health disorders in electronic healthcare record (EHR) notes changes over time due to updated disease definitions and clinical practice guidelines [10]. The terms used to describe mental disorders in clinical notes may acquire new definitions or shift meaning when their context changes, known as semantic drift. Such discontinuity in meaning between terms used in older clinical notes and new clinical notes can not only affect the accuracy of diagnostic and prognostic algorithms that were trained with historic data [11, 12] but also can impact the interpretation and classification of medical conditions [13, 14].

Semantic drift not only applies to diagnostic terms but also affects the other language used within these notes: This can be caused by variations in describing patient symptoms or by the use of acronyms.

This study sought to identify and measure semantic drift in the clinical notes of anxiety patients for two types of words: (a) terminology used by experts to diagnose anxiety, and (b) general clinical language. We used temporal word embeddings extracted from pediatric EHR clinical notes from 2009 to 2022. We measured semantic drift using methods to quantify temporal change in the word embeddings and evaluated the differences in semantic change between anxiety-related and general clinical terms.

## 2 Methods

### 2.1 Data Description

The Cincinnati Children’s Hospital Institutional Review Board approved this retrospective case-control study as STUDY2020-0942. As part of this study, we extracted EHR data from the Cincinnati Children’s Hospital Medical Center (CCHMC) Epic Link. This database has approximately 1.3 million unique patients seen at CCHMC between January 1, 2009, and March 31, 2022, with 63 million clinical notes. We included patients aged 0 to 25 in our pediatric population based on the definition provided by CCHMC. By including all data within these ages ranges, we gain a general understanding of how drift is affecting all notes in the corpus. While including age 0 is not conventional, it should not bias our results. For this study, we considered clinical notes only if they were created on or before the patient turned 25 years old; this exclusion left 61.5 million clinical notes available for analysis. Cases and controls were matched 1:1 using age and sex. CCHMC de-identified the data before sharing it with the authors by replacing names with individual IDs from 1 to 1.3 million.

### Preprocessing

Each clinical note contained a description of the patient’s visit. We preprocessed these notes by removing special characters, HTML tags, quotes, and brackets; replacing periods, tabs, and specialized line breaks with spaces; dropping stopwords; and removing PHI using Spacy NER library [15]. After these steps, we tokenized words from each clinical note by creating uni-grams for single words, bi-grams for common two-word phrases, and tri-grams for common three-word phrases. Figure **??** displays the inclusion criteria and data processing steps.

### 2.2 Diachronic Word Embedding

#### Word Embedding

Word embedding is a technique to represent words as dense vectors in a continuous vector space [16], where similar words have similar vector representations. We utilized word embedding to generate contextual vector representations of the words and the context in which a particular word was used in our collection of texts. Leveraging word embedding, we aimed to capture the contextual nuances of language evolution over time within our corpus of texts.

To achieve this, we devised a strategy to train individual Word2Vec [17] models in our dataset spanning from 2009 to 2022. This approach captures the shifting linguistic patterns and contextual variations in language usage across different years.

Each Word2Vec model was trained using the same parameters: a 100-dimensional word embedding, a window size of 5, a minimum count of 5, and a skip-gram with a negative sampling of 20. These parameters were chosen to be consistent with the literature [18–20]. We set the minimum count to 5 to allow for rare clinical words. We created 14 distinct Word2Vec models, each corresponding to a specific year in our dataset. Each Word2Vec model created a unique vector space where the words from the corresponding year were embedded. The vectors representing identical words across distinct models may exhibit dissimilar positions or orientations within their respective vector spaces.

#### Alignment of Word Embedding

The Word2Vec models were aligned following the methods of Hamilton et al. [20]. This alignment process was necessary because, upon completion of training, each model possessed its own unique vector space, making direct comparisons between models challenging. To address this challenge, each pair of models was systematically aligned to the previous year’s aligned model, ensuring a consistent transformation over time. This iterative alignment process enabled us to establish a coherent framework for comparing word embeddings across successive years, thereby facilitating the exploration of language evolution and semantic shifts.

We employed a word-to-word alignment method: Orthogonal Procrustes [21–23]. This method preserved the geometric properties while mapping vectors to the same spaces. This method also ensured that semantic similarities between words were preserved across different periods, enhancing the interpretability and reliability of our embeddings.

It’s important to note that this alignment method restricted our vocabulary to words that appeared at least once in every year’s model. As the alignment process aimed to find the best transformation from one year’s vector space to another, it relies on shared vocabulary across both years. Without this shared vocabulary, there is no basis for aligning the embeddings of words that do not appear in both years. This constraint helped maintain the coherence and meaningfulness of the alignment process, allowing for accurate comparisons and analyses of semantic relationships across different periods. The alignment process reduced our vocabulary size from 3.9 million unique words to 57,459 unique words (i.e., 1.5% of the total dataset).

### 2.3 Type of Word

To capture semantic change in expert-provided anxiety-related terminology and general EHR words, we then categorized the 57,459 words as *type* anxiety-related terms or *type* common EHR: this allowed us to assess how semantic drift in anxiety-related terms differed from semantic drift in common EHR terms.

#### Anxiety-Related Terms

We used a dictionary of words that were associated with at-risk anxiety diagnosis (AR). These words were collected from 19 mental health experts using the questions in Appendix Section A.1. We have 154 terms and phrases in the AR words type category, as seen in Appendix Table A.1.

#### Common EHR

The common EHR terms are words in the clinical notes that are not classified as AR. We do not further refine this category because it allows us to measure semantic drift in words common to pediatric clinical notes.

### 2.4 Measurements of Semantic Drift

Medical language changes over time as definitions of diagnoses are updated [13]. Capturing this change in language is important for applications that utilize longitudinal data. We measure this change over time through semantic drift.

We deployed methods to investigate semantic drift in anxiety-related expert language found in pediatric clinical notes and compared it to semantic drift in common language found in the same clinical notes. First, we measured semantic change using cosine distance. Second, we identified the semantic context using the neighborhood of a word. Finally, we deployed the laws of semantic change [20]. These laws provided two linguistic metrics for measuring semantic change: the law of conformity and the law of innovation. We further adapted the method to include a *type* variable specific to our corpus, as explained in section 2.2 [19]. The law of conformity was quantified using frequency, and the law of innovation was quantified using polysemy.

#### Measure of Semantic Change

Semantic change for an individual word between two years can be measured using the cosine distance between two aligned word vectors associated with the years. This metric measured the change of a word between two selected years and ranges between 0 and 1. A cosine distance of 0 means the vectors are completely the same, indicating no change in meaning. A cosine distance of 1 means the two vectors are complete opposites, indicating a maximal change in meaning. We used equation 1 to measure the semantic change of words in our dataset:

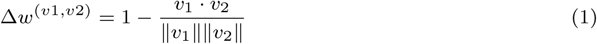

where Δ*w*^(*v*1*,v*2)^ represents the semantic change in a word for two different years, *v*1 represents the diachronic word embedding vector from the starting year, and *v*2 represents the diachronic word embedding vector from the ending year.

We provided an analysis to examine which AR terms and common EHR terms showed the greatest changes. Additionally, we analyzed how many terms fell within 0.25 quartiles of semantic change over time: this quantified the severity of year-wise semantic changes in the corpus. We defined the following quartiles for cosine difference: (1) 0.00 – 0.25: Very high similarity; (2) 0.26 – 0.50: High similarity; (3) 0.51 – 0.75: Moderate dissimilarity; (4) 0.76 – 1.00: High dissimilarity. Cosine distances were calculated using the SciPy library [24], words within the bottom two quartiles (0.00 - 0.50) were considered stable.

#### Measure of Semantic Neighborhood

Neighborhood semantics provided insight into the context of a word by finding the nearest K words using cosine similarity [25]: we created our neighborhood using the closest ten words to the word of interest. We applied this technique to each word of interest for every year in our dataset. Here, we investigated the local neighborhood of the AR word with the greatest average semantic change. Using Gensim to calculate cosine similarity [17], we compared words closely related to our word of interest.

Measuring the neighbors of a word in two different years shows how the context around a word changed from year to year. Semantically similar words were assumed to be found in similar contexts [26]. We quantified the change in a word’s context by describing the change in closely related words over time. It is important to note that when examining a word’s neighborhood, we included words from both the common EHR and the AR dictionaries to evaluate the neighborhood. A cosine similarity of 1 meant the two vectors were completely similar. We used equation 2 to measure the cosine similarity:

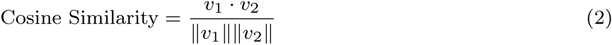

where Cosine Similarity measures the similarity between two vectors from two different years, *v*1 represents the diachronic word embedding vector from the starting year, and *v*2 represents the diachronic word embedding vector from the ending year.

#### Frequency

The law of conformity [20] can be used to frame our hypothesis about the relationship between semantic drift and how frequently a word is used; the more frequently a word is used, the slower it will change compared to words used infrequently. This law implies that because standard medical language is used at a high frequency, it should remain stable over time.

We measure this by calculating the relative word frequency for each word in our dataset for each year of interest and then dividing it by the total number of words in that year’s dataset.

#### Polysemy

The law of innovation [25] can be used to frame our hypothesis about the relationship between polysemy and semantic drift. Words used in many contexts, or polysemy, will change faster than words with fewer meanings. This law implies that ambiguous information inside clinical notes will have higher rates of semantic change over time. For example, it is not uncommon for clinical notes to contain ambiguous information, such as vague or subjective language and abbreviations. Vague language may include sentences such as, “Patient is doing well,” which is relative to the person writing the note and the person they are evaluating. Subjective language may include rating pain from “mild to severe”: a rating highly dependent on the person being asked.

Abbreviations can have multiple meanings depending on the healthcare provider or institution. One such example includes the abbreviation GAD, which may represent *generalized anxiety disorder* diagnosis or a tool used to access it named *generalized anxiety disorder-7* [27].

We measure polysemy by calculating the negative of the clustering coefficient: we used the Networkx package for extracting this metric [28]. The clustering coefficient is a measure used in network analysis to quantify the degree of interconnectedness or clustering of nodes in a network. By utilizing the negative clustering coefficient as a measure of polysemy, we aimed to quantify the extent to which words exhibited multiple meanings or senses based on their co-occurrence patterns within the dataset.

#### Quantifying Semantic Change

We can quantify the change in semantic meaning by modeling how a word’s rate of change is affected by word frequency and polysemy via a mixed linear model, shown in equation 3.

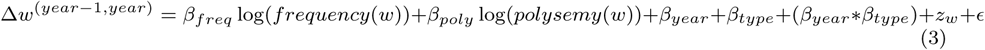

where Δ*w*^(^*^year−^*^1^*^,year^*^)^ is the log-normalized mean unit variance of semantic change, *β_freq_* represents the relative frequency of the word, *β_poly_* represents the polysemy, *β_year_* is the year of interest, *β_type_* is 1 for AR and 0 for common EHR, (*β_year_* ∗ *β_type_*) represents an interaction term between year and type, *z_w_* is the random effect that accounts for the word specific rate of change over repeated measures of time, and *ɛ* is an error term. In our equation, we included an interaction term to test for statistical differences for each year for AR words and common EHR words.

## 3 Results

### 3.1 Data

Our dataset included 61.5 million clinical notes and 3.9 million unique vocabulary words. The year 2014 had the most clinical documents, with almost 5.5 million clinical notes, and the year 2021 had the largest vocabulary size, with 969,843 unique words. Working within the common vocabulary space, our dataset was filtered to 57,459 words, which accounted for 1.5% of the total vocabulary size. Of the 154 AR terms, 60 appeared in our final dataset. Table 1 displays the number of documents available for each year and the individual years’ vocabulary size.

**Table 1.** Distribution of Number of Clinical Documents and Vocabulary Size across the years.

| Year | # Documents (%) |  | Vocabulary Size (%) |  |
| --- | --- | --- | --- | --- |
| 2009 | 1,633,364 | 2.7% | 284,080 | 7.2% |
| 2010 | 3,208,288 | 5.2% | 571,846 | 14.4% |
| 2011 | 4,398,965 | 7.1% | 657,691 | 16.6% |
| 2012 | 4,990,655 | 8.1% | 641,374 | 16.2% |
| 2013 | 5,141,910 | 8.3% | 698,865 | 17.6% |
| 2014 | 5,480,705 | 8.9% | 707,781 | 17.9% |
| 2015 | 4,757,635 | 7.7% | 649,381 | 16.4% |
| 2016 | 5,068,435 | 8.2% | 805,142 | 20.3% |
| 2017 | 5,080,136 | 8.2% | 836,113 | 21.1% |
| 2018 | 5,058,796 | 8.2% | 853,592 | 21.5% |
| 2019 | 5,221,043 | 8.5% | 891,642 | 22.5% |
| 2020 | 4,722,228 | 7.7% | 866,854 | 21.9% |
| 2021 | 5,430,950 | 8.8% | 969,843 | 24.5% |
| 2022 | 1,392,437 | 2.3% | 570,772 | 14.4% |
| **Total** | **61,585,547** |  | **3,961,421** |  |

### 3.2 Semantic Change

#### Type Evaluation

The AR word with the greatest change was *deviant* (average semantic change = 0.34), which is 6.7 times greater than the AR word with the least change (average semantic change *anxiety* = 0.05). The common EHR word with the greatest change was *OND* (average semantic change = 0.637): The common EHR word with the least change was *mom* (average semantic change = 0.03).

OND and ONS could be medical abbreviations, *OND* which has up to twenty different definitions, including *optic nerve disease, other neurological disorders, obese non-diabetic, and Ondansetron*, and *ONS* which has up to nine different definitions, including *other nervous system, optical nerve sheath, and optical nerves* [29]. Without context, we cannot determine which of these is the correct definition in our dataset. Another term observed in the dataset *nother*, as seen in Table 2, may be shorthand for “another” or a misspelling of “mother”. These three words may be used in multiple contexts in our dataset, corresponding to the moderate dissimilarity cosine score.

**Table 2.**
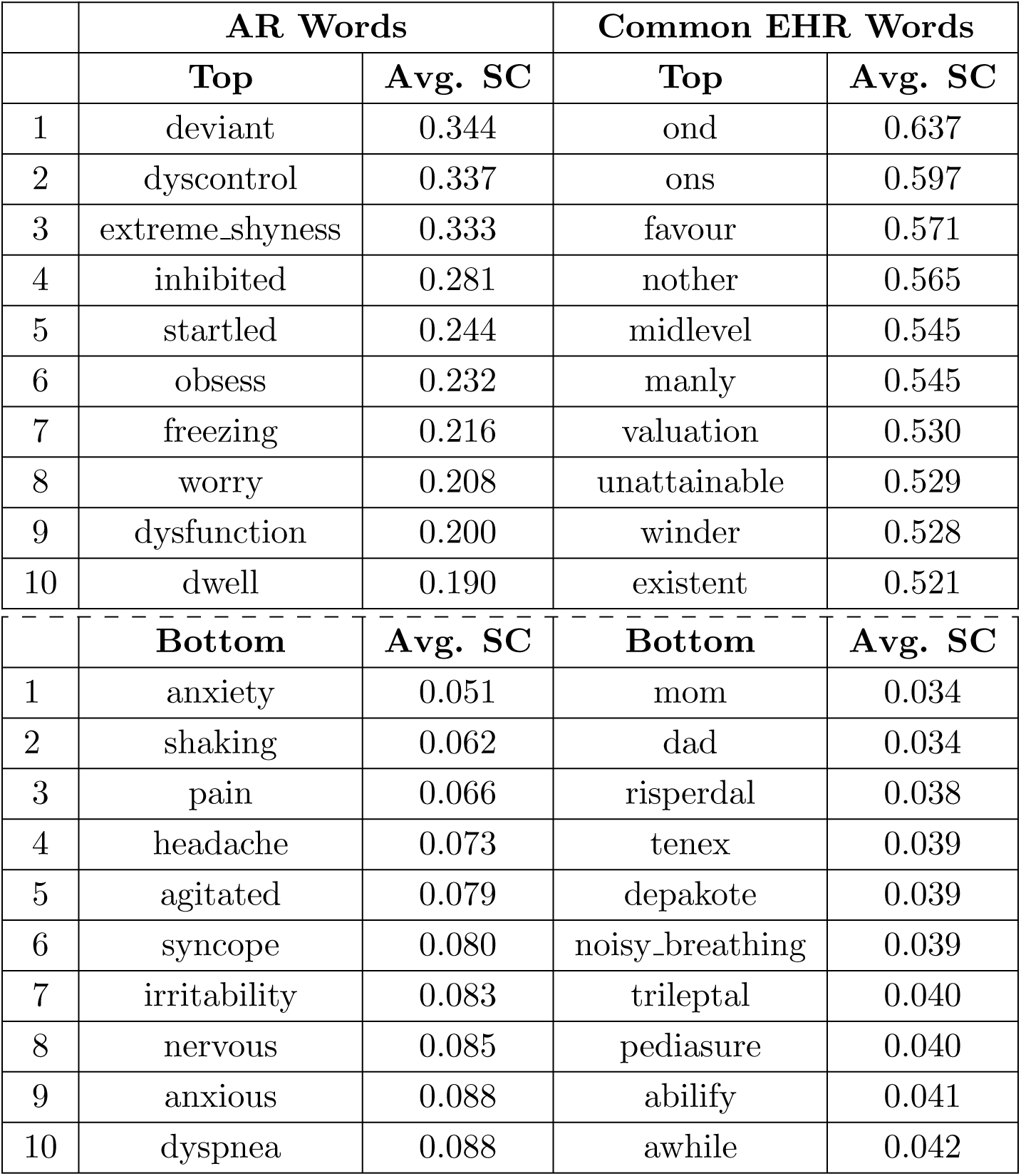
The Top 10 AR and common EHR words with the greatest change and the Bottom 10 AR and common EHR words with the least change. 0.00 – 0.25: Very high similarity; 0.26 – 0.50: High similarity; 0.51 – 0.75: Moderate dissimilarity; 0.76 – 1.00: High dissimilarity.

The average semantic drift of the highest-changing AR word was approximately half that of the highest-changing common EHR word (AR = 0.344; EHR = 0.637). The average change for the top 10 highest-changing AR words was less than that for the top 10 changing common EHR words (AR = 0.344 – 0.190; EHR = 0.637 – 0.521). For words with the lowest average semantic change, the change in the lowest AR word was 1.5 times greater than in the lowest common EHR word.

#### Quartiles of Semantic Change

Here we provide two comparisons of the number of words within the quartiles of semantic change: (1) AR words and (2) common EHR words. Table 3 displays semantic change calculated by year for all words in our dataset.

**Table 3.**
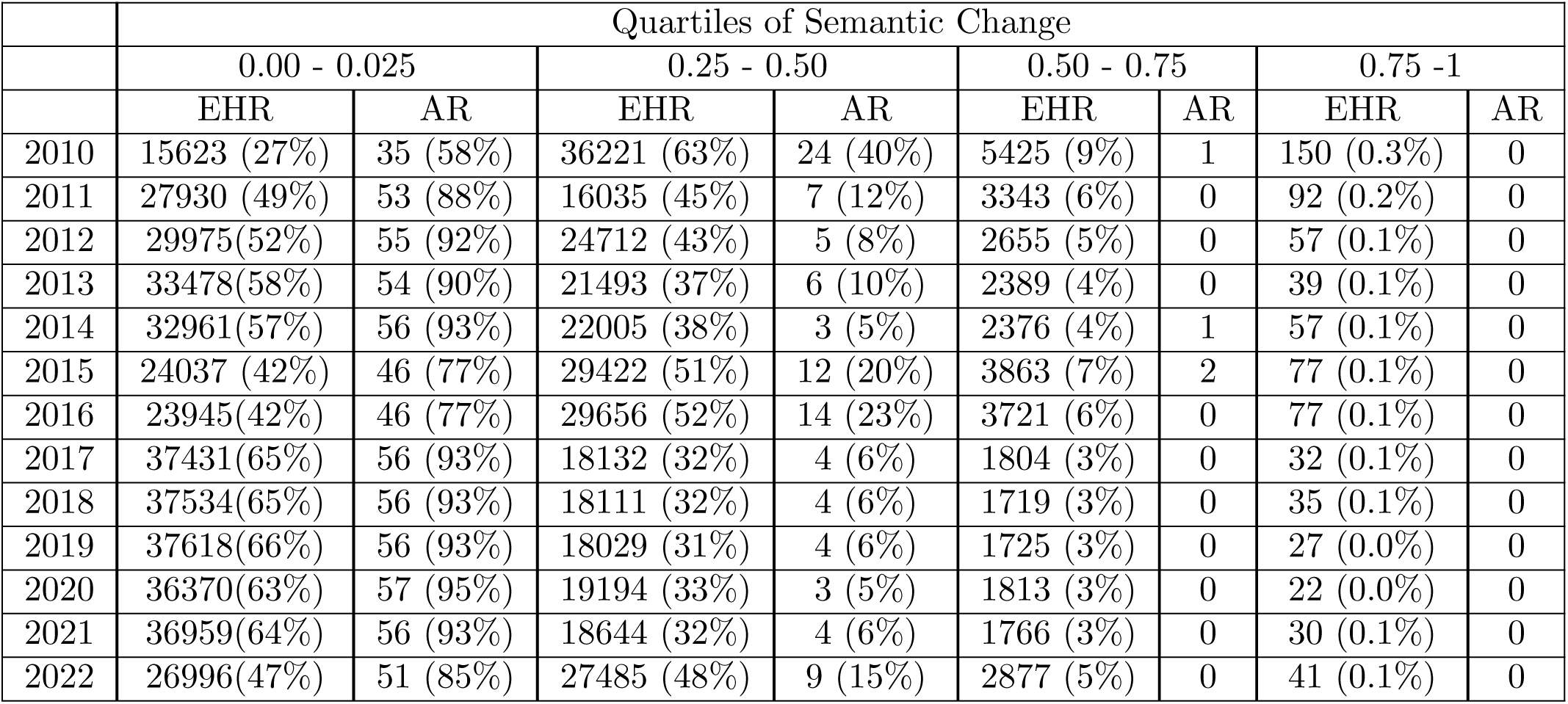
Number of words within 0.25 quartiles of semantic change from 0 to 1. 0.00 – 0.25: Very high similarity; 0.25 – 0.50: High similarity; 0.50 – 0.75: Moderate dissimilarity; 0.75 – 1.00: High dissimilarity.

Vocabulary Size % = Total Unique Words for Each Year / Total Unique Words. Total Percentages in the Vocabulary Size % Column may not sum to 100% given that unique words in each year will occur in other years.

For the AR data, 2020 had the highest number of words with very high similarity, while 2010 had the lowest (semantic change quartile 0.00 - 0.25: 2020 = 95%; 2010 = 58%). The year 2010 had the highest number of words with high similarity, while 2014 had the lowest (semantic change quartile 0.25 - 0.50: 2010 = 40%; 2014 = 5%).

Combining the quartiles for very highly similar and high similarity suggests that 98% – 99% of AR words in our dataset maintained a high degree of similarity throughout our dataset: this suggests that most AR words in our dataset are semantically stable.

For the common EHR words, 2019 had the highest number of words in the very highly similar category, while 2010 had the lowest number (semantic change quartile 0.00 - 0.25: 2019 = 66%; 2010 = 27%). The year 2010 had the highest number of words with high similarity, while 2019 had the lowest (semantic change quartile 0.25 - 0.50: 2010 = 63%; 2010 = 31%). Combining the quartiles for very highly similar and highly similar words suggested that 90% – 97% of words in our dataset maintained a high degree of similarity throughout our dataset, suggesting that the words in our dataset are semantically stable. The year 2010 had the highest number of words with moderate dissimilarity, while 2017 – 2021 had the lowest (semantic change quartile 0.50 - 0.75: 2010 = 9%; 2017 – 20212 = 3%). All years had fewer than 200 words in the high dissimilarity quartile (semantic change quartile 0.75 - 1.00: 0.1% – 0.3%). This demonstrates very few words in our vocabulary underwent a moderate to high semantic shift from their original context.

### 3.3 Neighborhood Semantics

We investigated the nearest neighbors of the AR word *deviant* (i.e., the word with the highest change reported) to gain insight into the changing context. Table 4 displays the top 10 nearest word neighbors for every year from 2009 – 2022 for the AR word *deviant*.

**Table 4.** Top 10 Neighbourhood Words for the word ‘deviant’ across the years Bold values represent words that have repeated more than one year Red words appear four times. Blue words appear three times. Forest Green words appear twice.

|  | AR Word – Deviant |
| --- | --- |
| Year | Top 10 Neighbors |
| 2009 | perpetuate, hinders, uncoordinated_movements, subscale., loading_phase, injuring_himself, sense_of_selfworth, mechanics, dynamics, eccentric_loading |
| 2010 | avoidant, histrionic, victimization, selfabsorbed, loudyelling, suggestible, **fantasy**, circuitous, personalities, devious |
| 2011 | **fantasy_world**, **grandiose**, **fantasy**, family, tends_to_obsess, problem, acts_strangely, creatures, anime, taken_advantage |
| 2012 | **provocative**, **bizarre**, hostile, stigmatizing, odd_prosody, pressured, paranoid, **grandiose**, apathetic, regressive |
| 2013 | revenge, implied, **confronted**, trust, believing, upsetting, trusted, people’s, trusting, realizing |
| 2014 | fueled, **provocative**, intimidated, offended, victimized, hypersexual_behaviors, selfmuiltating, being_teased, troubled, taunted |
| 2015 | concelament, indiscriminately, strongwilled, deliberately, distinguish, directionshe, biased, columbine, publicly, selfdepreciating |
| 2016 | revenege, malicious, adultlike, misperceive, negatively, enmeshed, indiscriminately, contrary, deliberately, overtly |
| 2017 | **unruly**, vulgar, **disrespect**, shouting, pouty, graphic, foul_language, **confronted**, **grandiose**, intimidating |
| 2018 | **bizarre**, **fixated**, pleasing, **fantasy_world**, severged, teenage, parenting_style, erratic, somewhat_chaotic, fascination |
| 2019 | overly, inappropriate, **bizarre**, natured, **fantasy**, obsessed, touchyfeely, contradict, portray, fascination |
| 2020 | **unruly**, intimidating, defying, stealing, paranoid, manipulative, inappropriateness, **shoplifting**, liar, stupid |
| 2021 | **unruly**, premeditated, remorse, rebellious, **disrespect**, reckless, **shoplifting**, malicious, hostility, disrespectful |
| 2022 | **bizarre**, menacing, blatant, **fixated**, retaliation, stalking, inappropriate_sexual, intrusive, reprimanded |

**Table 5.** Mixed Linear Model Results: Variable and corresponding coefficient for their effect on semantic change. positive coefficient score corresponds to a faster rate of change and negative coefficient score corresponds to a slower rate of change

| Variable | Laws of Semantic Change |  |  |
| --- | --- | --- | --- |
|  | Coefficient | P-Value | Confidence Int. 95% |
| Intercept | 2.206 | $p < 0.000$ | (2.188, 2.224) |
| Frequency $\beta_{freq}$ | -0.04 | $p < 0.000$ | (-0.043, -0.039) |
| Polysemy $\beta_{poly}$ | 0.630 | $p < 0.000$ | (0.620, 0.641) |
| Type w/out Interaction $\beta_{type}$ | -0.179 | $p < 0.000$ | (-0.307, -0.051) |
| 2010 | -2.022 | $p < 0.000$ | (-2.028, -2.015) |
| 2011 | -2.374 | $p < 0.000$ | (-2.382, -2.367) |
| 2012 | -2.432 | $p < 0.000$ | (-2.440, -2.424) |
| 2013 | -2.565 | $p < 0.000$ | (-2.574, -2.557) |
| 2014 | -2.521 | $p < 0.000$ | (-2.530, -2.512) |
| 2015 | -2.181 | $p < 0.000$ | (-2.191, -2.171) |
| 2016 | -2.154 | $p < 0.000$ | (-2.166, -2.144) |
| 2017 | -2.679 | $p < 0.000$ | (-2.691, -2.668) |
| 2018 | -2.683 | $p < 0.000$ | (-2.694, -2.672) |
| 2019 | -2.685 | $p < 0.000$ | (-2.697, -2.694) |
| 2020 | -2.619 | $p < 0.000$ | (-2.631, -2.608) |
| 2021 | -2.631 | $p < 0.000$ | (-2.644, -2.620) |
| 2022 | -2.285 | $p < 0.000$ | (-2.296, -2.275) |

The results revealed that 10 words appeared in multiple years, of which several may be specific to age.

It is worth noting that some words in the table, although spelled differently, refer to the same meaning; an example of this is *tends to obsess* and *obsessed*. We treated these words as separate instances and tracked them individually throughout the dataset.

We provide an additional evaluation of the top 10 neighbors for the AR word with the least change, *anxiety*, for every year in our dataset in Appendix Section A.2. The results revealed that the majority of the words in the neighborhoods were repeated.

### 3.4 Testing the Laws of Semantic Change

Here, we report the coefficients for variables *Word Frequency, Polysemy, Type, and Year*, allowing us to test the statistical laws of semantic changed proposed by [20]. A negative coefficient implies a slow rate of semantic change and a positive coefficient implies a fast rate of semantic change. The mixed linear model allowed us to quantify the relationship between frequency, polysemy, word type, and semantic change.

We found that words which occurred frequently in our dataset had a slow rate of change (Coef = -0.04): words with higher polysemy scores changed fast (Coef = 0.630). On average, AR words changed slower than EHR words (Coef = -0.179).

We hypothesized that the semantic differences may vary over our 14-year observation period and used a (Type * Year) interaction to assess differences in the effect of word type over time. Semantic differences were the least in 2010, with AR words having insignificant differences (Coef = -.09; P-value *<* 0.276): The largest differences were in 2017 (Coef = -0.523). For the remaining years, the rate of change for AR words was significantly slower than for EHR words (Coefs = -0.275 – -0.523). The magnitude of this difference was reduced in 2015, 2016, and 2022. These results from 2015 and 2016 may indicate changes in the DSM criteria during this time. However, this hypothesis was not directly tested: 2022 may be due to the limited data within this year. To validate these results, we examined the frequency and polysemy values for the AR words and common EHR words with the highest and lowest semantic change, as well as the highest and lowest frequency words in the dataset.

For words with the least semantic change, the AR word *anxiety* appeared 1,150,779 times, while the common EHR word *mom* appeared 5,704,186 times. For words with the greatest semantic change, the AR word *deviant* appeared 559 times, while *ond* appeared 499 times. For the most frequent words in the dataset, the AR word *pain* appeared 6,957,928 times, while the common EHR word *patient* appeared 18,263,934 times. For the least frequent words in the dataset, the AR word *dysfunction* appeared 534 times, while the common EHR word *conjunctival* appeared 474 times. The frequency of AR word *anxiety* was only 20% of the frequency of common EHR word *mom*, and the frequency of AR word *pain* was only 38% of the frequency of common EHR word *patient*. These results indicate that less frequently used words changed faster than more frequently used words and validated the type variable.

Polysemy values for the common EHR word *ond* were greater than polysemy values for the AR word *deviant* (ond polysemy range = (-0.09, -0.01); deviant polysemy range = (-0.31, -0.04)). Polysemy values for AR word *anxiety* were higher than polysemy values for common EHR word *mom* (anxiety polysemy range = (-0.37, -0.15); mom polysemy range = (-0.33,-0.20)). These results are reflected in the average semantic change values.

Figuredisplays the rate of change for AR and common EHR words from the MLM. This figure demonstrates that the AR word group changes slower than common EHR words (i.e., the blue and orange lines).

For words with the least semantic change, the AR word *anxiety* appeared 1,150,779 times, while the common EHR word *mom* appeared 5,704,186 times. For words with the greatest semantic change, the AR word *deviant* appeared 559 times, while *ond* appeared 499 times. For the most frequent words in the dataset, the AR word *pain* appeared 6,957,928 times, while the common EHR word *patient* appeared 18,263,934 times. For the least frequent words in the dataset, the AR word *dysfunction* appeared 534 times, while the common EHR word *conjunctival* appeared 474 times. The frequency of AR word *anxiety* was only 20% of the frequency of common EHR word *mom*, and the frequency of AR word *pain* was only 38% of the frequency of common EHR word *patient*. These results indicate that less frequently used words changed faster than more frequently used words and validated the type variable.

Polysemy values for the common EHR word *ond* were greater than polysemy values for the AR word *deviant* (ond polysemy range = (-0.09, -0.01); deviant polysemy range = (-0.31, -0.04)). Polysemy values for AR word *anxiety* were higher than polysemy values for common EHR word *mom* (anxiety polysemy range = (-0.37, -0.15); mom polysemy range = (-0.33,-0.20)). These results are reflected in the average semantic change values.

Figuredisplays the rate of change for AR and common EHR words from the MLM. This figure demonstrates that the AR word group changes slower than common EHR words (i.e., the blue and orange lines).

### 3.5 Summary

As seen from the results, at least 98% of the AR words and at least 90% of the common EHR words remained highly similar in semantics. We found that our dataset conformed to the Laws of Semantic Change. Frequent words changing slowly in our dataset implies that common medical terminology in AR and common EHR words is stable.

Alternatively, words with higher polysemy scores, such as medical abbreviations or other ambiguous terms, changed quickly. The top common EHR word, OND, reflects this, showing the highest average semantic shift. AR words changed faster than common EHR words; however, the overall average change for AR was slower than common EHR.

## 4 Discussion

The NLP-based methodology described here can be applied to any clinical decision support system (CDSS) that utilizes longitudinal text data. Evolving healthcare practices that update definitions and guidelines facilitate the need to evaluate language over time within clinical notes, as CDSS must be robust to temporal shifts in the data. Deploying NLP-based techniques that capture semantic drift as it occurs can reveal when data has changed contextually too much from the original meaning: Discontinuity between terms used to describe diagnoses or treatments can decrease data quality and may impact downstream CDSS that leverage NLP and provide unreliable predictions [3]. These techniques can provide insight into when to stop using data from certain years or when retraining models with more current data is necessary. Utilizing 63 million clinical notes collected from CCHMC’s pediatric population, we identified and quantified semantic drift in the expert-provided anxiety-related terminology and common EHR language.

Stable language can be used to anticipate clinical events and patient trajectories. Trajectory modeling utilizes longitudinal data to predict patient outcomes. Hence, the stability in clinical language ensures that meaningful insights can be extracted from clinical notes. While both word sets maintained a high degree of similarity throughout our dataset, we found that AR words changed faster than general EHR language.

Notably, frequently used words from both categories remained relatively stable over time. Drug names, as seen in Table 2, showed little to to no semantic drift due to their standardized nature and consistent usage in clinical contexts. However, abbreviations such as *ond* posed a challenge due to their high polysemy scores, potentially introducing ambiguity in CDSS interpretation. The context in which abbreviations occurred must be accurately captured to mitigate the risk of errors influencing the downstream models. Our study did not explicitly limit our clinical note population to anxiety patients; therefore, the interpretation of these findings may be impacted when considering an anxiety-specific population. Additionally, we did not consider clinical notes based on age groups. Considering that age may reveal considerably different neighborhoods of AR words and may highlight interesting trends in years: anxiety terminology is likely to appear different for a group of children compared to teenagers. In future work, we will examine anxiety-specific clinical notes and clinical notes belonging to specific ages.

The quantity of anxiety-related terms included in this study was limited compared to the general EHR words. While our original AR terms included 154, only 60 appeared in our final dataset. This highlights the need for a more comprehensive list, including abbreviations. In addition to the AR terms being limited, the shared vocabulary space was limited. As seen in Table 1, we did not consider most of the unique vocabulary words in our dataset. The alignment process can limit additional insights gained from these words. For example, we may miss words such as “COVID-19”, “pandemic”, or “Coronavirus” in 2020, 2021, and 2022 and their effect on our anxiety-related terms [30]. We believe that using 2009 as the starting year in our dataset further limited our vocabulary: This was the first year in which the EHR was adopted at CCHMC. Data may likely be missing or incomplete for this year due to the scanning of medical documents, late adoption of the EHR system, and errors in initial familiarization with the system. As a result, using 2009 as the starting year immediately led to excluding words that appeared in subsequent years but not in 2009. Future work should consider ways to include out-of-vocabulary words when examining semantic drift for more insightful evaluations. Additionally, many of the common EHR words with the greatest change are misspellings or uncommon spellings in American English, highlighting a limitation in our analysis. Addressing this in future work will improve the insights gleaned from semantic drift evaluations.

Understanding the semantic drift around anxiety diagnostic criteria in clinical notes can reduce opportunities for outdated diagnoses and treatment plans for downstream predictive modeling [31]. The effect of updating ICD and DSM codes was not fully considered here; however, we completed a brief ingestion into Asperger’s [14]. We completed a brief investigation into the word Asperger’s: Asperger’s was chosen because it is now included under Autism in the change from DSM-4 to DSM-5 [11]. We present the results in Appendix Section A.4. Future work should examine semantic drift related to DSM codes, specifically in years before and after changes.

## 5 Conclusion

This study investigated semantic drift in expert-provided anxiety-related terminology compared to general vocabulary found in clinical notes. Using multiple methods, we investigated semantic drift evaluating year-wise semantic change, the semantic neighborhood, and applying two linguistic measures of semantic drift. We found that words in our dataset remained semantically stable. Our future work includes applying insights gained here to anxiety trajectory modeling.

## Data Availability

Data cannot be shared publicly because it contains personally identifiable information and would be protected health information as such, we are not authorized to make our dataset publicly available. This data was acquired and used under an approved IRB protocol from Cincinnati Children's Hospital Medical Center. Data are available from the Cincinnati Children's Hospital Medical Center Institutional Data Access / Ethics Committee for researchers who meet the criteria for access to confidential data.

## Acknowledgments

This work was supported by Cincinnati Children’s Hospital Medical Center under Strategic Partnership Projects agreement NFE-21-08617. This abstract has been authored by UT-Battelle, LLC under Contract No. DE-AC05-00OR22725 with the U.S. Department of Energy. This research used resources of the Oak Ridge Leadership Computing Facility, which is a DOE Office of Science User Facility supported under Contract DEAC05-00OR22725.

## Appendix

### A.1 Expert Defined Terms

The anxiety-related (AR) terms for this study were defined by 19 CCHMC mental health experts answering three questions:

1. Anxiety Similar Words: Besides the term “anxiety,” what other words, phrases, or terms would you use in the medical records to designate the presence or severity of anxiety?

2. Anxiety at Risk Words: What characteristics or indicators indicate that the patient may have an anxiety diagnosis in the future?

3. Anxiety Key Phrases: What characteristics or indicators make you sure that the patient does have anxiety?

These three questions yielded 154 terms, as seen in Appendix Table A.1.

### A.2 Extended Neighbors

We evaluated our dataset’s top ten neighbors for every year to further quantify semantic drift for the AR words with the least change, *anxiety*.

Table A.2 displays the AR word anxiety results. The results reveal eighteen words appear in multiple years. Additional words mean the same thing but are spelled differently; examples include *anxieties, anxietymood*, and *anxious*.

These results highlight the need to refine the vocabulary tokenization pre-processing step further so that these words can be considered one word in our dataset.

### A.3 Mixed Linear Model

Here, we report the coefficients for variables *Law of Conformity, Law of Innovation, Type, and Year* for interacting the year and type coefficients. We found the rate of change for polysemy and frequency do not change (frequency Coef = -0.04; polysemy Coef = -0.179); refer to Table A.3.

### A.4 Asperberg’s Syndrome

Diagnostic criteria that change with simple language swaps such as *any* of these symptoms to *all* of these symptoms will change the inclusion and exclusion criteria of a diagnosis after an update [32]. We completed a brief investigation into ERH common word *Asperger’s* from 2009 to 2014: Asperger’s does not appear in our dataset after 2014. Asperger’s was chosen because it is now included under Autism in the change from DSM-4 to DSM-5 [11].

**Table A.1.**
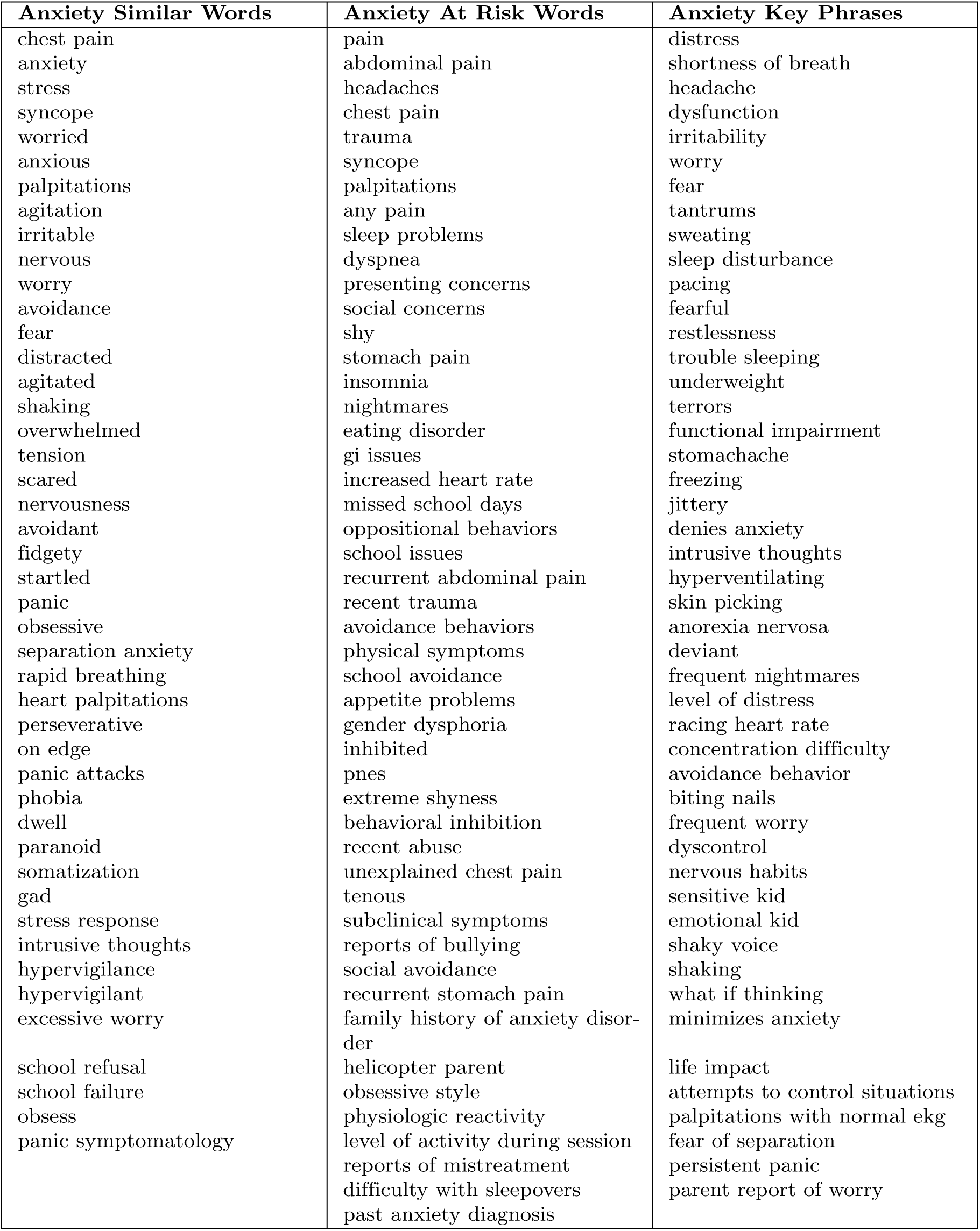
Full list of anxiety-related (AR) terms provided by the CCHMC mental health experts.

**Table A.2.**
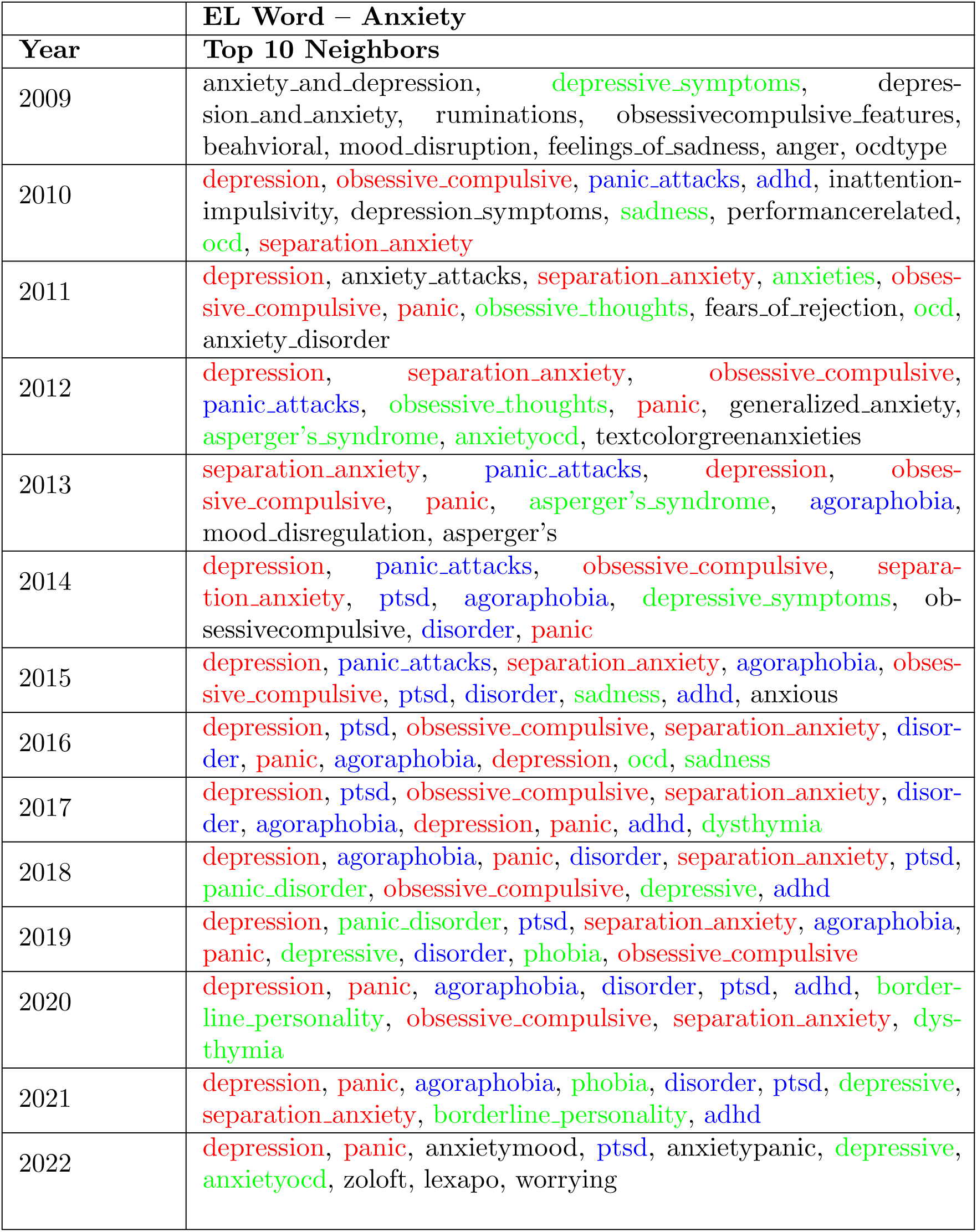
Top 10 Neighborhood for AR word *anxiety* across all years in our dataset. Red words appear 11 to 15 times. Blue words appear 5 to 10 times. Forest Green words appear less than 5 times.

**Table A.3.** Mixed Linear Model Results: Variable and corresponding coefficient for their effect on semantic change. positive coefficient score corresponds to a faster rate of change and negative coefficient score corresponds to a slower rate of change

| Variable | Coef. | P-Value | Conf. Int. 95% | Variable | Coef. | P-Value | Conf. Int. 95% |
| --- | --- | --- | --- | --- | --- | --- | --- |
| Intercept | 2.206 | p < 0.000 | (2.187, 2.224) |  |  |  |  |
| Relative Frequency | -0.04 | p < 0.000 | (-0.043, -0.039) |  |  |  |  |
| Polysemy | 0.63 | p < 0.000 | (0.620, 0.641) |  |  |  |  |
| Type | 0.174 | p < 0.043 | (0.005, 0.343) |  |  |  |  |
| 2010 | -2.022 | p < 0.000 | (-2.028, -2.015) | Type * 2010 | -0.09 | p < 0.276 | (-0.251, 0.072) |
| 2011 | -2.374 | p < 0.000 | (-2.381, -2.366) | Type * 2011 | -0.478 | p < 0.000 | (-0.639, -0.317) |
| 2012 | -2.432 | p < 0.000 | (-2.440, -2.424) | Type * 2012 | -0.501 | p < 0.000 | (-0.663, -0.340) |
| 2013 | -2.565 | p < 0.000 | (-2.573, -2.556) | Type * 2013 | -0.466 | p < 0.000 | (-0.628, -0.305) |
| 2014 | -2.521 | p < 0.000 | (-2.530, -2.511) | Type * 2014 | -0.394 | p < 0.000 | (-0.555, -0.232) |
| 2015 | -2.181 | p < 0.000 | (-2.191, -2.171) | Type * 2015 | -0.238 | p < 0.002 | (-0.399, -0.077) |
| 2016 | -2.154 | p < 0.000 | (-2.165, -2.143) | Type * 2016 | -0.258 | p < 0.004 | (-0.419, -0.096) |
| 2017 | -2.679 | p < 0.000 | (-2.690, -2.668) | Type * 2017 | -0.523 | p < 0.000 | (-0.685, -0.362) |
| 2018 | -2.683 | p < 0.000 | (-2.694, -2.671) | Type * 2018 | -0.437 | p < 0.000 | (-0.598, -0.276) |
| 2019 | -2.685 | p < 0.000 | (-2.697, -2.674) | Type * 2019 | -0.395 | p < 0.000 | (-0.556, -0.234) |
| 2020 | -2.619 | p < 0.000 | (-2.631, -2.608) | Type * 2020 | -0.386 | p < 0.000 | (-0.547, -0.224) |
| 2021 | -2.631 | p < 0.000 | (-2.643, -2.620) | Type * 2021 | -0.507 | p < 0.000 | (-0.668, -0.345) |
| 2022 | -2.285 | p < 0.000 | (-2.295, -2.275) | Type * 2022 | -0.275 | p < 0.001 | (-0.436, -0.114) |

**Table A.4.**
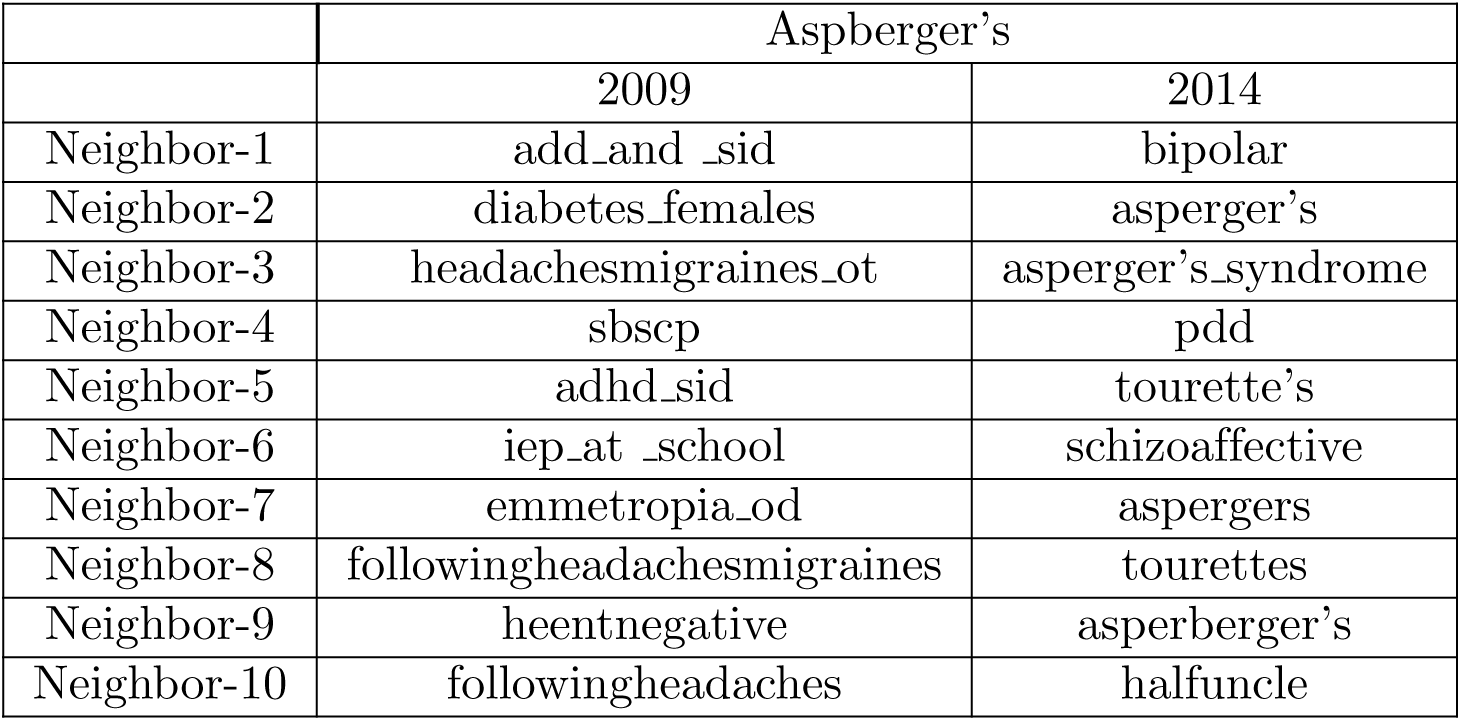
Top 10 Neighborhood for EHR word *Aspberger’s* from 2009 to 2014.

---

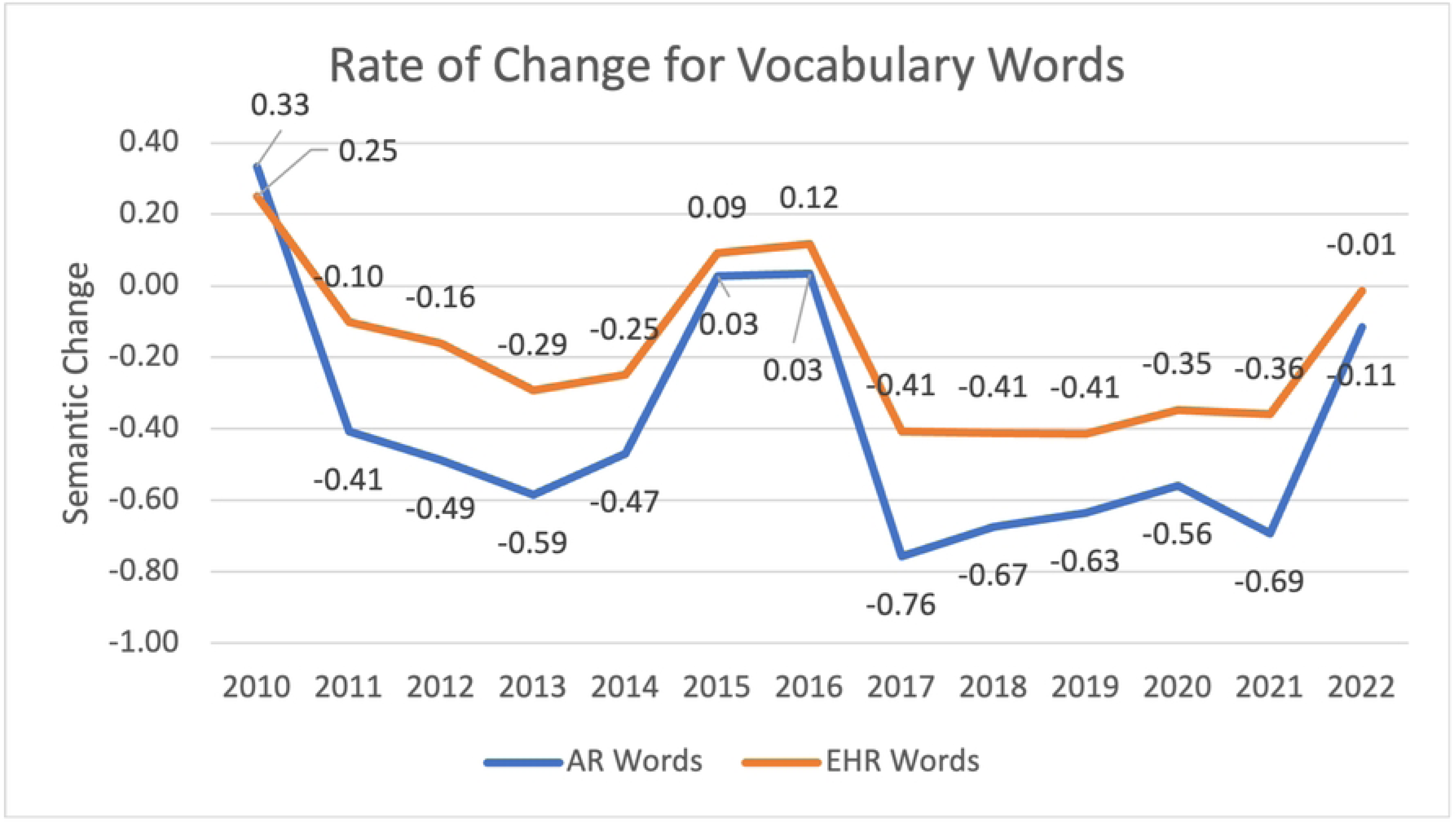

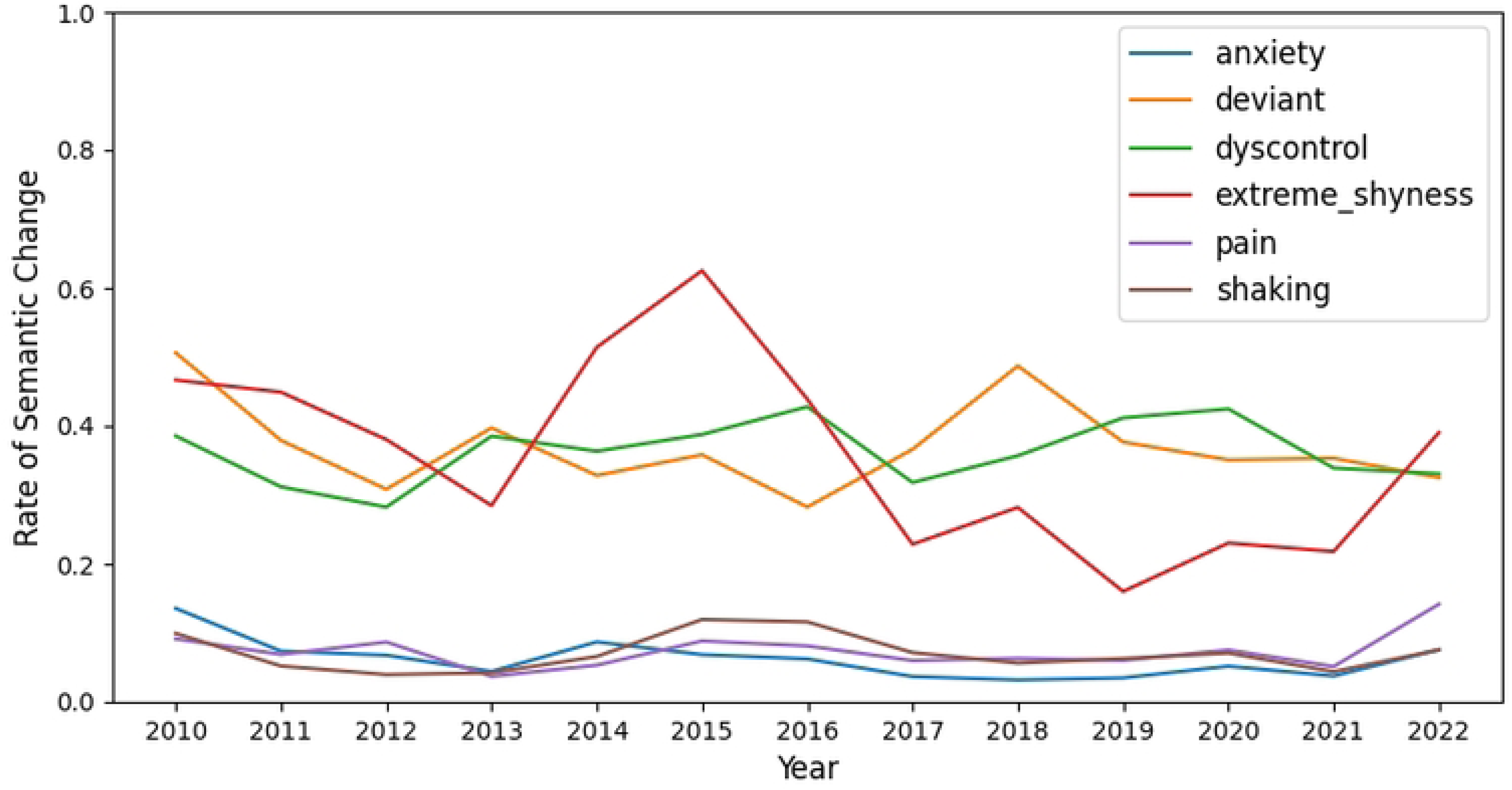

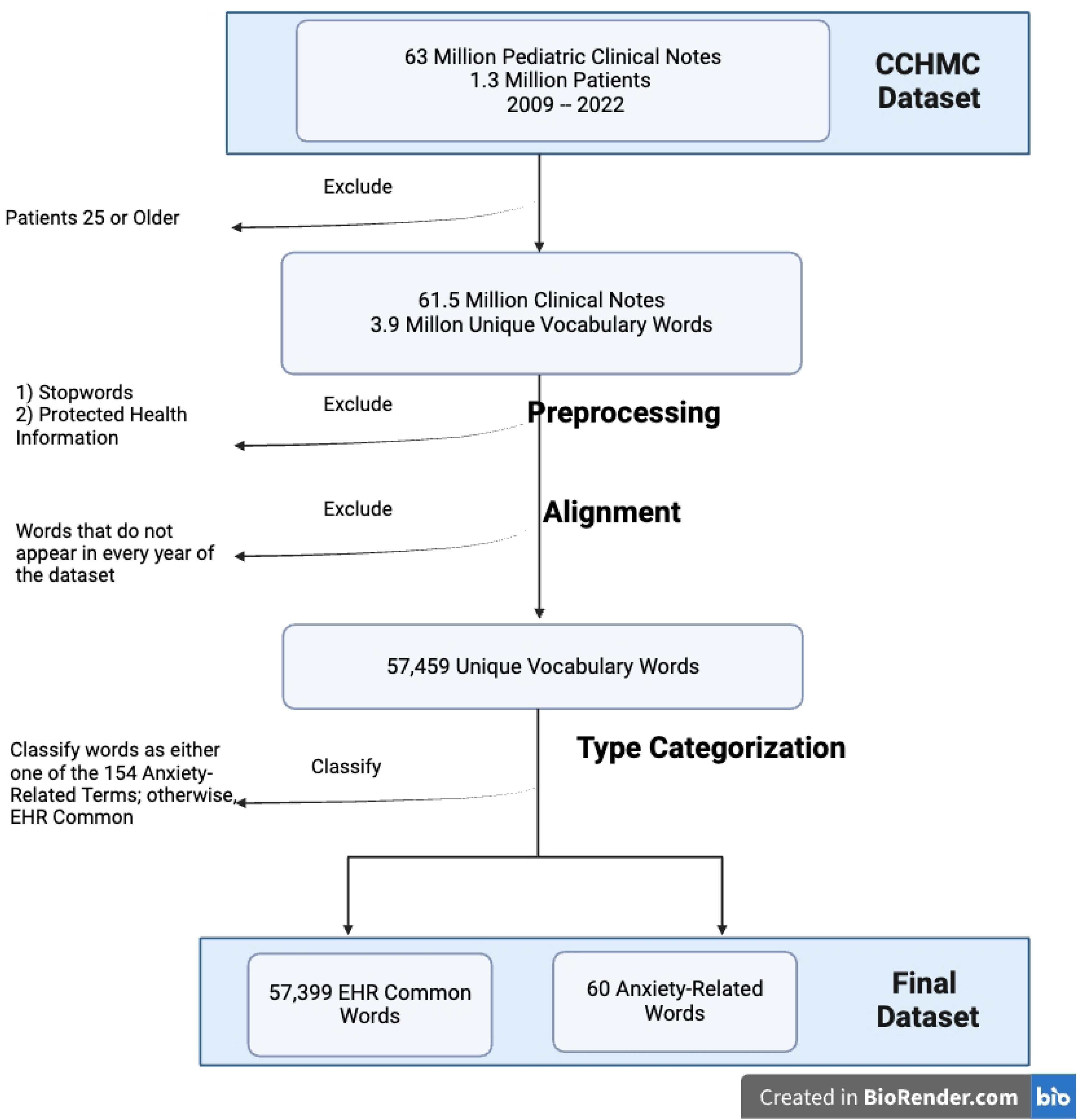

